# Rare variants in *PRKCI* cause Van der Woude syndrome and other features of peridermopathy

**DOI:** 10.1101/2025.01.17.25320742

**Authors:** Kelsey Robinson, Sunil K. Singh, Rachel B Walkup, Dorelle V. Fawwal, Wasiu Lanre Adeyemo, Terri H. Beaty, Azeez Butali, Carmen J. Buxó, Wendy K. Chung, David J. Cutler, Michael P. Epstein, Azeez Fashina, Brooklynn Gasser, Lord JJ Gowans, Jacqueline T. Hecht, Lina Moreno Uribe, Daryl A. Scott, Gary M. Shaw, Mary Ann Thomas, Seth M. Weinberg, Harrison Brand, Mary L. Marazita, Robert J Lipinski, Jeffrey C. Murray, Robert A. Cornell, Elizabeth J. Leslie-Clarkson

## Abstract

Van der Woude syndrome (VWS) is an autosomal dominant disorder characterized by lower lip pits and orofacial clefts (OFCs). With a prevalence of approximately 1 in 35,000 live births, it is the most common form of syndromic clefting and may account for ∼2% of all OFCs. The majority of VWS is attributed to genetic variants in *IRF6* (∼70%) or *GRHL3* (∼5%), leaving up to 25% of individuals with VWS without a molecular diagnosis. Both IRF6 and GRHL3 function in a transcriptional regulatory network governing differentiation of periderm, a single layer of epithelial cells that prevents pathological adhesions during palatogenesis. Disruption of this layer results in a spectrum of phenotypes ranging from lip pits and OFCs to severe pterygia and other congenital anomalies that can be incompatible with life. Understanding the mechanisms of peridermopathies is vital in improving health outcomes for affected individuals. We reasoned that genes encoding additional members of the periderm gene regulatory network, including kinases acting upstream of IRF6 (i.e., atypical protein kinase C family members, RIPK4, and CHUK), are candidates to harbor variants resulting in VWS. Consistent with this prediction, we identified 6 *de novo* variants (DNs) and 11 rare variants in *PRKCI*, an atypical protein kinase C, in 17 individuals with clinical features consistent with syndromic OFCs and peridermopathies. Of the identified DNs, 4 were identical p.(Asn383Ser) variants in unrelated individuals with syndromic OFCs, indicating a likely hotspot mutation. We also performed functional validation of 12 variants using the enveloping layer in zebrafish embryos, a structure analogous to the periderm. Three patient-specific alleles (p.Arg130His, p.(Asn383Ser), and p.Leu385Phe) were confirmed to be loss-of-function variants. In summary, we identified *PRKCI* as a novel causal gene for VWS and syndromic OFC with other features of peridermopathies.

## Introduction

Van der Woude syndrome (VWS, MIM:119300) is an autosomal dominant disorder that occurs in approximately 1 in 35,000 live births^1,2^. It is commonly characterized by bilateral lower lip pits and an orofacial cleft (OFC)^3,4^. Approximately 70% of VWS is caused by heterozygous variants in *IRF6*, including whole gene deletions^5,6^ and structural variants^7^, missense variants, and truncating variants^8–10^. Another ∼5% of VWS is caused by missense or truncating variants in *GRHL3*^11^, leaving up to 25% of cases unsolved. While all families with VWS may present with lip pits, a cleft lip with or without a cleft palate (CL/P), and/or a cleft palate only (CP), there is some evidence for a genotype-phenotype correlation. Individuals with *GRHL3* variants are more likely to have CP compared to those with *IRF6* variants^11^. However, unsolved VWS families span the phenotypic spectrum of OFCs and lip pits. Gene discovery for Mendelian conditions, including VWS, is made easier when large, affected families are available, regions of significant linkage have been identified, and there is genetic homogeneity. In the absence of these, as is the case for many unsolved VWS families, gene discovery can be difficult unless unidentified causal genes can be easily tied to associated biological pathways.

VWS belongs to a group of conditions now referred to as peridermopathies, which result from disruption of the periderm. The periderm is a single epithelial cell layer lining the oral cavity and other structures during development, and functions to prevent pathologic adhesions in early embryogenesis^12^. Disturbance of this cell layer can lead to clinical features ranging from mild to incompatible with life. Peridermopathies include popliteal pterygium syndrome (PPS, MIM:119500), caused by heterozygous variants in *IRF6* that result in lower lip pits, OFCs, sygnathia, pterygia (webbing) of the lower limbs, syndactyly, and urogenital abnormalities^13,14^.

Peridermopathies with more severe phenotypic manifestations include Bartsocas-Papas syndrome (BPS; MIM 263650) and Cocoon syndrome (MIM: 613630) caused by homozygous or compound heterozygous variants in *RIPK4*^15,16^ and *CHUK* (also called *IKK1* and *IKKa*)^17^, respectively.

The genes implicated in peridermopathies belong to a complex transcriptional regulatory network (TRN) and signaling cascade that control cell differentiation. In mammalian embryos, the periderm is derived from basal keratinocytes^12^, whereas in zebrafish the periderm analog— the enveloping layer (EVL)—is derived from blastomeres^18^. Yet, there are many similarities between the TRNs of mammals and zebrafish, and much of what is known about this network comes from research in both model systems. IRF6 and GRHL3 are conserved transcription factors that play key roles in periderm development in both mammalian^11,19–23^ and zebrafish^24–27^ embryos. IRF6 directly activates expression of *GRHL3*^26^ leading to periderm differentiation.

IRF6 is phosphorylated and activated by RIPK4^28,29^, which also phosphorylates CHUK and IKKb^30^, resulting in activation of NF-kB signaling^31^. The phenotype of *Chuk/IKKa* loss of function mutants closely resembles that of *Irf6* loss of function mutants in both mouse^32^ and zebrafish^33^, linking this kinase to periderm development. The activation of RIPK4 at the top of this TRN is not well elucidated, but RIPK4 interacts with protein kinase C (PKC) and both overexpression and knockdown of RIPK4 alters the keratinocyte differentiation response to PKC activation^28,34^. In addition, the atypical PKCs (aPKCs iota, *PRKCI*, and zeta, *PRKCZ*) are pivotal in determining cell fate of the superficial ectoderm in frog embryos^35,36^.

Individuals in this cohort with VWS and without pathogenic variants in *IRF6* or *GRHL3* also lack pathogenic variants in *RIPK4* or *CHUK.* Because VWS shares phenotypes with the conditions associated with this network of genes, other genes upstream or downstream of *IRF6* are excellent candidates to harbor variants resulting in VWS. Here we report that variants of *PRKCI*, which encodes for the atypical protein kinase C iota (PKCι), are causal for a range of syndromic orofacial clefts including VWS and other clinical features of peridermopathies. We identified exome-wide enrichment of *de novo* variants (DNs) in *PRKCI* associated with syndromic CP, including a recurrent variant, c.1148A>G, p.(Asn383Ser) found in four individuals and several rare, protein-altering, predicted damaging variants. We tested the functional effects of 12 variants using a zebrafish model and found that 3 of these, including p.(Asn383Ser), result in loss of function, consistent with the development of a peridermopathy and other phenotypes in humans.

## Materials and Methods

### Study cohort

VWS: 17 families diagnosed with VWS with no known pathogenic variant in *IRF6* or *GRHL3* based on bidirectional Sanger sequencing were included in this study. Institutional review board approval was obtained for each local recruitment center and coordinating centers including University of Iowa, University of Pittsburgh, and Emory University.

OFCs: We also analyzed two large OFC cohorts: one ascertained primarily on CP, which will be referred to as CPseq, and the other ascertained on any OFC as part of the Gabriella Miller Kids First (GMKF) Research Consortium. For CPSeq, a total of 435 trios with CP were recruited from diverse ancestral backgrounds including European (recruited from Spain, Turkey, Hungary, United States), Latin American (Puerto Rico, Argentina), Asian (China, Singapore, Taiwan, the Philippines), and African (Nigeria, Ghana). For GMKF, case-parent trios originate from three ancestral groups: 376 trios of European (recruited from sites in the United States, Argentina, Turkey, Hungary, and Spain), 267 trios from Medellin, Colombia, and 116 trios from Taiwan. In total, there were 1,511 affected individuals from both CPSeq and GMKF broken down into 608 CP, 897 CL/P, and 3 unknown OFC type. Phenotyping, recruitment, and details of local ethical approval were previously described for CPSeq^37^ and GMKF^38^. All recruitment was approved by local review boards and/or coordinating centers at the University of Iowa, the University of Pittsburgh, Johns Hopkins University, or Emory University.

### Sample preparation and whole genome sequencing (WGS)

VWS and CPSeq cohorts: WGS was performed at the Center for Inherited Disease Research (CIDR) at Johns Hopkins University (Baltimore, MD). Alignment, variant calling, and quality control was performed using the DRAGEN Germline v3.7.5 pipeline on the Illumina BaseSpace Sequence Hub platform, producing single sample VCF files. Details on the full pipeline for sequencing have been published in Robinson et al^37^.

DNs were called using a separate pipeline than the full cohort multisample VCF. First, the DRAGEN 3.7.5 aligner and variant caller was used to generate gVCF files for each trio. The gVCF files along with a pedigree file for each trio were provided as input to the DRAGEN 3.7.5 joint caller, resulting in a trio specific VCF file including tags for potential *de novo* variants.

Genotypes were set to missing if GQ<20 or read depth <10. To be considered a DN, variants had to have a quality score of 30, DQ>2, and parental genotypes had to be confirmed homozygous reference (0/0), pass all filtering steps, and have an allele balance (AB) ratio of <0.05. Only *de novo* variants with MAF of <0.5% in gnomAD exomes v2.1.1 or gnomAD v3.1.2 were retained.

GMKF: WGS was performed from blood samples in the majority of cases, but saliva was used in cases when blood was unobtainable. Sequencing for European samples was carried out by the McDonnell Genome Institute (MGI) the Washington University School of Medicine (St. Louis, MO) followed by alignment to hg38 and variant calling at the GMKF Data Resource Center at the Children’s Hospital of Philadelphia. Sequencing for Colombian and Taiwanese samples was carried out by the Broad Institute, with alignment to hg38 and variant calling by GATK pipelines^39–41^. Additional details on alignment and workflow used to harmonize these datasets have been published in Mukhopadhyay et al^42^.

### Variant filtering and annotation

Variants with a passing filter flag were included. Next, we removed variants aligning outside of the standard chromosomes (1-22, X, Y) and with a minor allele count (MAC) of <1. Genotypes with GQ<20 or read depths of <10 were set to missing, and sites with missingness values of >10% were then removed. Sample-level quality control metrics included transition/transversion (Ts/Tv) ratio, silent/replacement rate, and heterozygous/homozygous ratio; outlier samples with values outside of 3 standard deviations from the cohort mean were discarded. Samples with high missing data (>5% missing) were removed.

All variants were annotated with Annovar (version 201707)^43^. Variants with a MAF of <0.5% in any population in either gnomAD exomes v2.1.1 or gnomAD v3.1.2 were considered rare for this analysis. We retained protein-altering variants, removing any annotated as synonymous. We estimated predicted pathogenicity with the *in silico* tools SIFT^44^, PolyPhen-2^45^, and AlphaMissense^46^.

### DN enrichment

Enrichment of DNs in *PRKCI* was calculated using the ‘DenovolyzeByGene’ function of the R package ‘DenovolyzeR’ (version 0.2.0)^47^. As 572 genes had at least 1 DN in the 475 CP trios, we established significance at p-values less than 8.74 x10^-5^ (Bonferroni correction for 572 genes). Exome-wide significance is reached for p-values less than 1.30x10^-6^.

### Protein 3D structure modelling

*In silico* modeling of *PRKCI* variants was conducted in PyMOL using PDB 1ZRZ. Variants were modeled using a backbone-independent rotamer and measured using PyMOL Wizard Measurement tool.

### Zebrafish

All the procedures carried out in this project involving zebrafish were approved by the Institutional Animal Care and Use Committee (IACUC) at the University of Washington, Seattle and follow NIH guidelines. Embryos from zebrafish wild-type AB strain were used for all the experiments. Embryos were grown at 28.5°C and staged according to the standard protocol^48^.

### ORF cloning and reverse transcription

Human PRKCI reference (with and without -CAAX sequence at 3’ end. C = cysteine, A = aliphatic amino acid; X = any amino acid) were synthesized and cloned in pCS2+ vector by GenScript (https://www.genscript.com/). *PRKCI* variants were generated by 1-bp site-directed mutagenesis in CAAX-version of reference gene sequence. Reference and variant-containing mRNAs were synthesized from 1µg linearized plasmids by using mMESSAGE mMACHINE™ SP6 Transcription Kit (Invitrogen; cat# AM1340). mRNA products were treated with DNase (Invitrogen™ DNA-free™ DNA Removal Kit; cat# AM1906) and purified using RNA Clean & Concentrator™-25 kit (Zymo Research; cat# R1017).

### Zebrafish studies

Zebrafish embryos were injected at 1-2 cell stage with samples containing approximately 200 pg mRNAs encoding reference or variant PRKCI. Injected embryos were kept in embryo medium (5.03 mM NaCl, 0.17 mM KCl, 0.33 mM CaCl_2_, 0.33 mM MgSO_4_, 0.1% Methylene blue) at 28.5°C until harvested for different assays.

### Whole mount *in situ* hybridization and quantitative real-time PCR

Antisense RNA probes for zebrafish *krt4* were synthesized by using primers 5′- TAAGACCCTCAACAACCGCT-3′ and 5′- TAATACGACTCACTATAGGGCTACCGTATCCTGACCCACC-3′^49^ and DIG RNA Labeling Kit (Roche; cat# 11175025910). Approximately 10 injected embryos per group were collected at 50-percent epiboly stage (5.5 hpf). The Whole mount *in situ* hybridization procedure was performed as described by Thisse and Thisse, 2008. Fixed *in situ* hybridized embryos were washed 3 times with 1x phosphate buffer saline (PBS) and soaked in 35-percent Sucrose solution for 4 hours. Embryos were stirred gently in Tissue-Tek optimal cutting temperature (OCT) embedding medium (Sakura Finetek; cat# 4583) to remove addition sucrose solution. Next, embryos were transferred to a cryomold filled with fresh OCT medium and frozen in ethanol with dry ice pellets. Twenty-micron sections were prepared from frozen specimen blocks using a Leica CM 1850 cryostat. Digital images of whole mount embryos and cryo-sections were captured using a K3C camera attached to Leica MZFL stereoscope

Approximately 40 mRNA-injected Shield-stage (6-hpf) embryos per group were processed for total RNA extraction using RNAqueous™ Total RNA Isolation Kit (Invitrogen; cat# AM1912). cDNA was prepared from approximately 1µg DNase treated total RNA using High-Capacity cDNA Reverse Transcription Kit (Applied Biosystems; cat# 4368814). Zebrafish *krt4* gene expression was measured in 10-fold diluted cDNA samples using iTaq Universal SYBR Green Supermix (Bio-Rad; cat# 1725121) on CFX Connect Real-Time PCR Detection System (Bio-Rad; cat# 1855201). Fold-change in *krt4* gene expression was calculated by using the 2^−ΔΔCT^ comparative CT method^50^ and zebrafish *actb1* as reference gene. Primers are listed in supplementary table **S1**.

### PRKC inhibitor treatment

The un-injected and *PRKCI* mRNA injected embryos were incubated at 28.5°C with 2 micromolar PRKC inhibitor (PKC_ζ_ Pseudosubstrate Inhibitor, Myristoylated – Sigma Aldrich; cat# 539624) in 0.5x Ringer’s solution with 0.01% DMSO. Embryos were photographed and scored for the rupture phenotype at approximately 4 hpf.

### PRKCI Western Blot

Approximately 40 zebrafish embryos (4 hpf) from every sample group were dechorionated and deyolked following the protocol by S. O’Shea and M. Westerfield^51^. Deyolked embryos were lysed and homogenized in ice-cold RIPA buffer containing protease inhibitors (Roche, cOmplete Mini) at 4 oC overnight with gentle shaking. The tissue lysate was centrifuged at 15,000 rpm for 10 minutes. The supernatant was quantified for the protein content using Pierce™ BCA Protein Assay Kit (Thermo Scientific; Cat# 23227). Approximately 6 μg of protein lysate was prepared in loading buffer (1x Laemmli sample buffer (Bio-Rad; Cat# 1610747), 2.5% 2-mercaptoethanol (Sigma Aldrich; Cat# 63689), 2M Urea), incubated at RT for 15 minutes, and then loaded onto a 10% SDS-polyacrylamide gel (Bio-Rad #4568034). Protein was transferred to polyvinylidene fluoride (PVDF) membranes (Thermo Scientific #88520). The membrane was blocked with 5- percent BSA in TBS buffer (Sigma Aldrich; Cat# A8022) and incubated overnight with PKC iota Antibody (Novus Biologicals; Cat# NBP1-84959) and GAPDH Antibody (Cell Signaling Technology #2118S) in TBS-T buffer. The membrane was washed three times with TBS-T buffer and incubated with horseradish peroxidase-conjugated goat anti-rabbit secondary antibody (Thermo Scientific; Cat# 32460) for 1 hour at room temperature. Finally, the membrane was washed three times with TBS-T buffer and developed using SuperSignal West Dura Extended Duration Substrate (Thermo Scientific #34075) and imaged on iBright CL1500 Imaging System.

### Mouse Studies

Mouse studies were performed in strict accordance with recommendations from the National Institutes of Health’s “Guide for the Care and Use of Laboratory Animals.” The animal care protocol for animals used in this study was approved by the University of Wisconsin-Madison School of Veterinary Medicine Institutional Animal Care and Use Committee (protocol no. V005396). Wild-type C57BL/6J mice were purchased from The Jackson Laboratory and housed in ventilated cages maintained at 22°C ± 2°C and 30%–70% humidity on a 12-hour dark cycle. Mice were provided with Irradiated Soy Protein-Free Extruded Rodent Diet (catalog no. 2920x; Envigo Teklad Global) until day of copulation plug after which pregnant dams received Irradiated Teklad Global 19% Protein Extruded Rodent Diet (catalog no. 2919; Envigo Teklad Global). Timed matings were achieved by placing one or two nulliparous female mice with a singly housed male mouse for 1-2 hours and then checking for copulation plugs. The beginning of the mating period was designated as gestational day 0 (GD0), and pregnancy was confirmed during gestation by assessing weight gain as previously described^52^. Dams were euthanized via carbon dioxide inhalation followed by cervical dislocation between GD11-GD14 ± 1 hour for embryo collection or GD15 ± 2 hours for fetal collection.

For *in situ* hybridization (ISH) assays, embryos at GD11, GD14, and GD15 were removed from the uterine horn in phosphate buffered saline (PBS) and fixed in 4% paraformaldehyde in PBS for 18 hours. Embryonic and fetal samples then underwent graded dehydration into 100% methanol (1:3, 1:1, 3:1, v/v) for storage at -20 °C. ISH riboprobes were designed using IDT PrimerQuest and their specificity was confirmed utilizing National Center for Biotechnology Information Primer Basic Local Alignment Search Tool (NCBI Primer-BLAST) (Table S2).

Whole-mount ISH for *Prkci* was performed as previously described^53,54^. Subsequently, tissues were embedded in 4% agarose and cut into sections using a vibrating microtome (50 μM for GD11, 100 μM for GD14 and GD15). Images of both whole-mount and sectioned tissues were captured using a MicroPublisher 5.0 camera (QImaging) mounted on an Olympus SZX-10 stereomicroscope.

Another cohort of embryos was used for quantitative gene expression analysis at GD11 and GD14. Embryos were collected and micro-dissected in PBS and enzymatic separation of ectoderm and mesenchyme from dissected maxillary processes (GD11) and palatal shelf tissues (GD14) was performed as described previously^55,56^. Total RNA was isolated from pooled tissue samples (from 6 embryos for GD11 samples and 3 embryos for GD14 samples) using the QIAGEN RNeasy Mini Kit (catalog no. 74104) with on-column DNase I digestion as per the standard protocol. A total of 100 ng RNA was used for cDNA synthesis using the Promega GoScript Reverse Transcriptase kit (Catalog no. A5000). Real-time qPCR primers (Table S2) were designed using IDT PrimerQuest and their target specificity was confirmed using the NCBI BLAST. Quantitative real-time PCR was performed using the Bio-Rad SSoFast EvaGreen Supermix (catalog no. 1725202) on a Bio-Rad CFX-Duet Real-Time PCR Detection

System. *Gapdh* was used as the housekeeping gene, and the expression of *Prkci* relative to that of *Gapdh* was calculated using the 2^−ΔΔCt^ method^50^. Paired, two-tailed t-tests were used to compare RNA expression in mouse embryonic tissues.

## Results

### *PRKCI* is enriched for *de novo* variants in VWS and OFC phenotypes

We performed whole genome sequencing (WGS) on 17 families diagnosed with VWS but negative for pathogenic variants in *IRF6* or *GRHL3* confirmed with bidirectional Sanger sequencing of both genes. This included 9 complete trios for which *de novo* variants (DNs) were called and evaluated. We found an exome-wide significant enrichment for DNs in *PRKCI* (p=8.56x10^-8^), a gene moderately constrained to both loss of function variation (LOEUF in gnomad v4.1.0=0.66) and missense variation (gnomad v4.1.0 Z-score = 3.07). Remarkably, this enrichment was the result of two individuals with identical DNs in *PRCKI* (NM_002740: c.1148A>G, p.[Asn383Ser]) and the same phenotype (lip pits with cleft soft palate). None of the other individuals with VWS harbored variants in *PRKCI*.

Lip pits are incompletely penetrant in VWS^57^, and those affected can present with any OFC. Therefore, we expanded our search to all rare variants (MAF <0.5%) in a cohort of 1,491 OFC case parent trios ascertained primarily on OFC but without restriction to additional clinical features (598 CP, 888 CL/P, and 3 unknown OFC type). We identified a third individual with a DN in *PRKCI* (NM_002740: c.407A>G, p.[Tyr136Cys]), who had a cleft soft palate as well as abnormal lip morphology, though had not received a diagnosis of lip pits or VWS. Additionally, there were 10 protein-altering, rare, inherited variants in 3 CP and 7 CLP probands.

Simultaneously, we queried clinical databases, such as GeneMatcher^58^ in which individuals are not ascertained on an OFC. In these databases, there were 59 rare, protein-altering variants with phenotypic information available for 25 individuals. Of these, 4 people had OFCs with DNs identified in 3 of them. Strikingly, two of these DNs were additional p.(Asn383Ser) variants. One individual was reported to have CP (subtype unspecified), global hypotonia, developmental delays, and autism spectrum disorder, while the second was reported to have a syndromic CL and CP with a prominent forehead and moderate intellectual disability. The third identified DN was just two amino acids downstream (c.1155A>C, p.Leu385Phe), reported in an individual with phenotypes consistent with a more severe peridermopathy: cleft hard and soft palate, sygnathia (adhesions between the upper and lower jaw), ankyloblepharon (adhesions between the upper and lower eyelids), elbow and knee contractures, as well as an atrial septal defect.

In total we compiled 72 rare, protein-altering variants in *PRKCI* (**Supplemental Table 3**) with phenotype information available for 38 probands (**Figure 1**, **Table 1**). In individuals with OFCs, there were 14 unique variants found in 17 individuals, with 6 DNs and 11 inherited variants. We did not see any clear clustering of variants based on protein domain, although 10 of 14 variants were within the large catalytic region (which makes up 61% of the protein).

**Figure 1:**
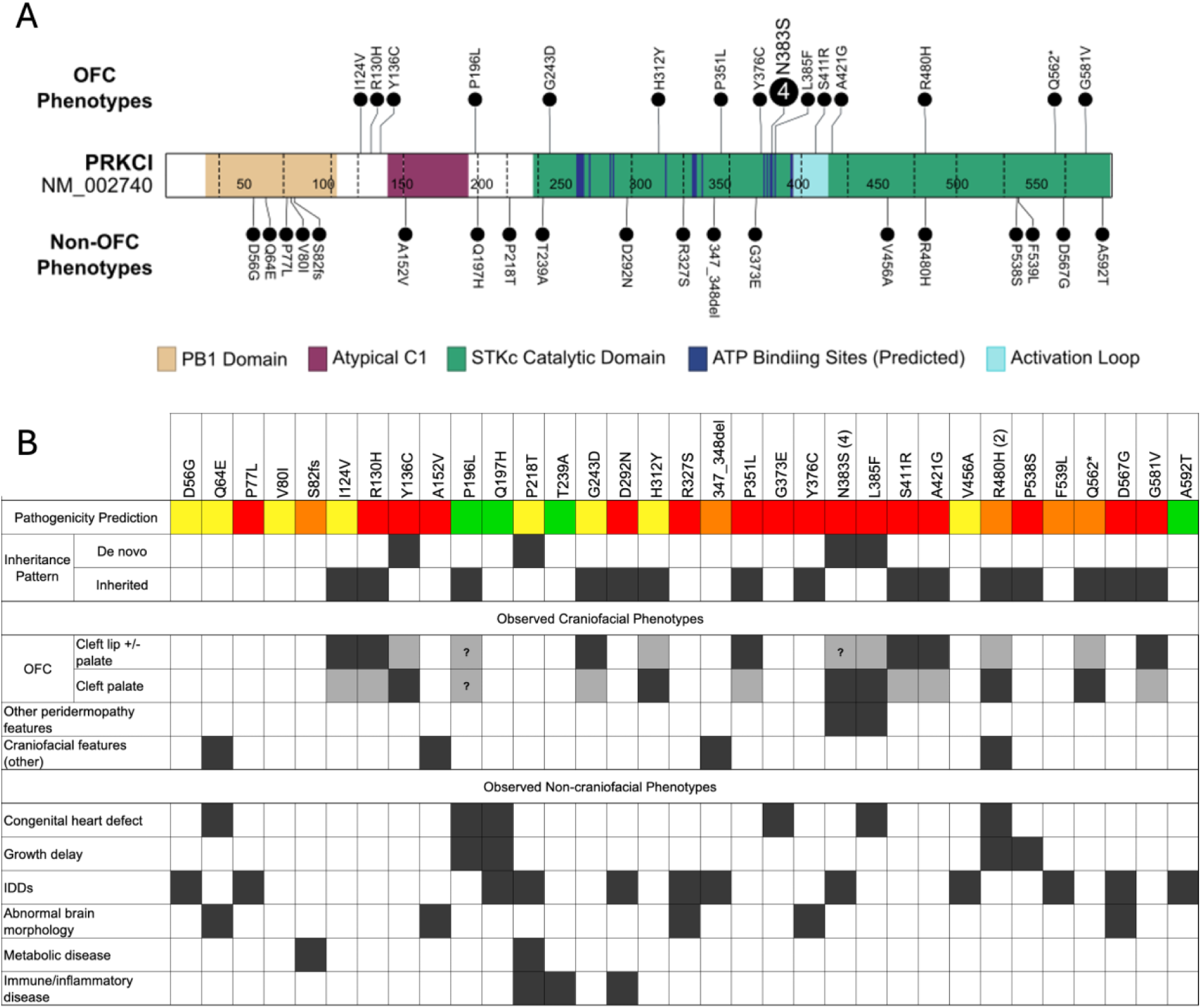
PRKCI structure, variant distribution, and proband phenotype descriptions. A) Lollipop plot of rare *PRKCI* variants in individuals with OFCs (top) and without OFCs (bottom) across the linear structure. The number of variants per site is indicated by the number in the circle, where a solid circle indicates a single variant. B) Phenotype and variant description grid for all variants shown in 1A. Pathogenicity predictions: green = predicted deleterious by 0 methods, yellow = predicted deleterious by 1 method, orange = predicted deleterious by 2 methods, red = predicted deleterious by 3 methods (algorithms used include SIFT, PolyPhen2, and AlphaMissense).

**Table 1:**
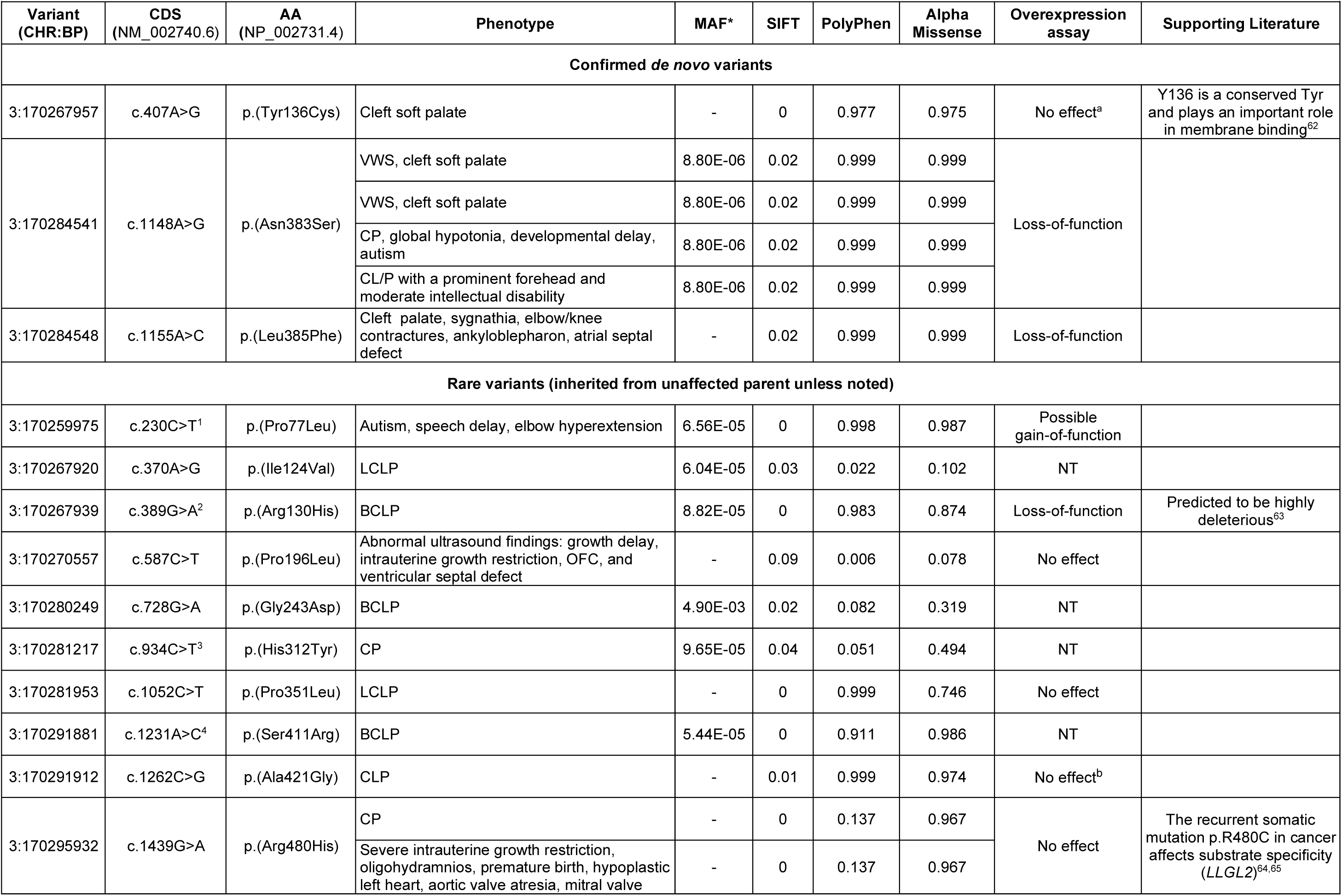
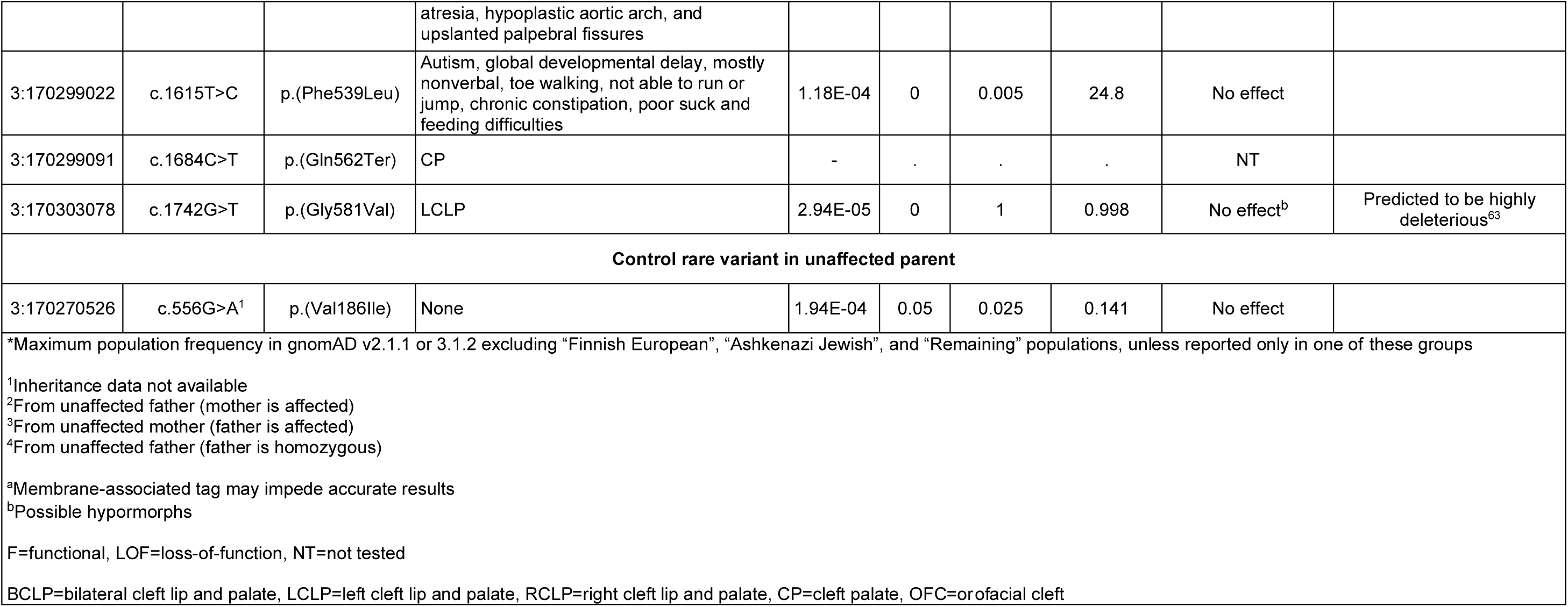
DNs and rare variants in *PRKCI* from all individuals with OFCs and/or functionally tested alleles.

However, both the p.(Asn383Ser) and p.Leu385Phe DNs are at predicted ATP-binding sites with the catalytic region^59,60^ which may directly interfere with kinase function^61^.

Given compelling genetic evidence for a role of *PRKCI* in peridermopathy-related phenotypes, we investigated its biological relevance in embryonic development.

### *PRKCI* is expressed in palatal tissue

*PRKCI* encodes atypical protein kinase C (aPKC) iota (PKCι), a serine/threonine protein kinase. Unlike conventional protein kinase C, aPKCs are Ca^2+^ and diacylglycerol independent. PKCι has three protein domains (**Figure 1A**). The Phox/Bem 1 (PB1) domain is important for guiding cell polarity^66^, the atypical C1 domain is considered part of the regulatory region^67^, and the protein kinase catalytic domain^66,67^ is responsible for catalytic activity. Other functions associated with PKCι have largely been studied in the context of cancer^65^, finding a variety of roles in cell differentiation, migration, and proliferation^68^.

*PRKCI* is expressed relatively ubiquitously, though expression appears dependent upon developmental time point and cell type. In bulk RNA sequencing of human craniofacial tissue^69^, *PRKCI* is expressed throughout Carnegie Stages (CS) 13-22 with a general trend downward over time **(Figure 2A**). At CS20, a single nucleus RNA sequencing dataset (also from craniofacial tissue)^69^ illustrates this ubiquitous expression, though *PRKCI* expression is strongest in the ectoderm cell cluster, which gives rise to the periderm (**Figure 2B**). Previous work of *in situ* hybridization in mice has shown *Prkci* expression throughout development in multiple tissues including regions of the brain, olfactory epithelium, liver, lungs, heart, stomach, and kidneys^70^. However, as this work did not focus on craniofacial development, we performed *in situ* staining for *Prkci* at several key stages of palatogenesis: fusion of the medial and maxillary processes (gestational day (GD), GD11), vertical outgrowth of palatal shelves (GD14), and subsequent horizontal outgrowth and fusion of the palate (GD15). Whole-mount and sectional images demonstrated *Prkc*i expression in the mesenchyme and ectoderm of the developing maxillary processes and palatal shelves during palatogenesis (**Figure 3A-C**). Subsequent quantitative analysis demonstrated that *Prkci* expression is significantly higher in the ectoderm compared to the mesenchyme at both GD11 and GD14 (**Figure 3D**). The ectodermal expression of *Prkci* was lower at GD14 compared to GD11, though the relative mesenchymal expression remained comparable at both timepoints.

**Figure 2:**
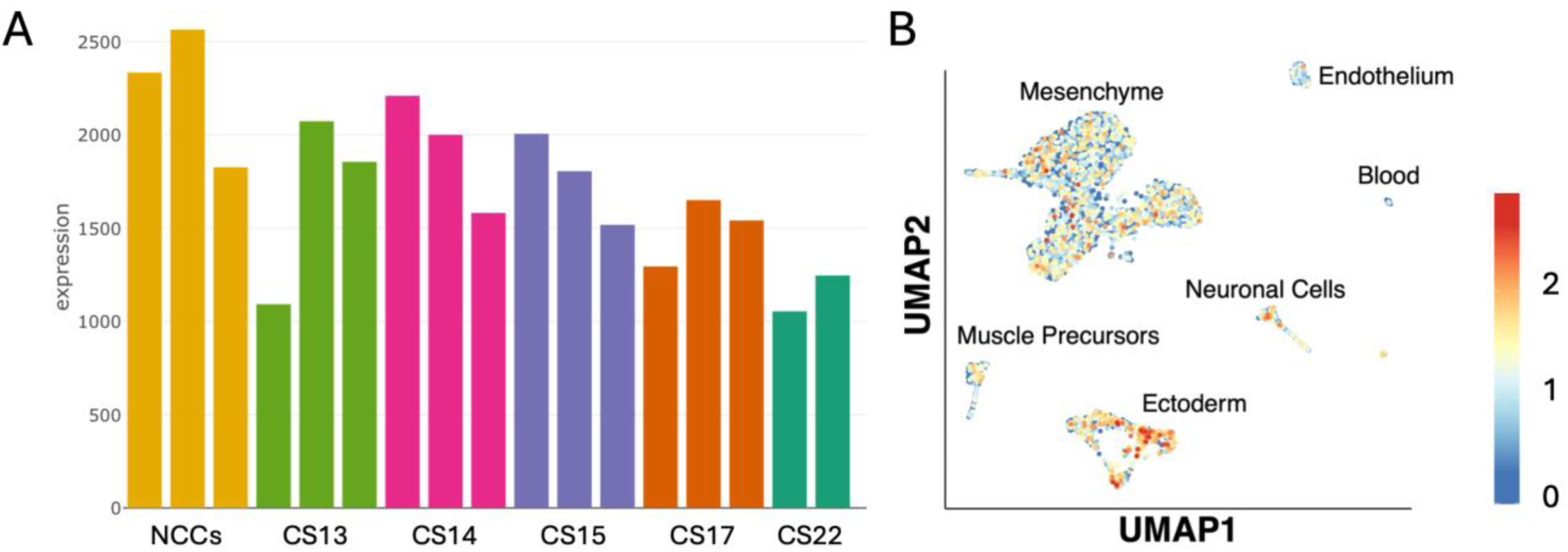
PRKCI is expressed in human craniofacial tissue during early embryogenesis. A) PRKCI expression in bulk RNA sequencing data for neural crest cells (NCC) and human craniofacial tissue from Carnegie Stages (CS) 13,14,15,17, and 22. B) PRKCI expression is highest in the ectoderm cell cluster in a dataset of single nucleus RNA sequencing from human craniofacial tissue at CS20.

**Figure 3:**
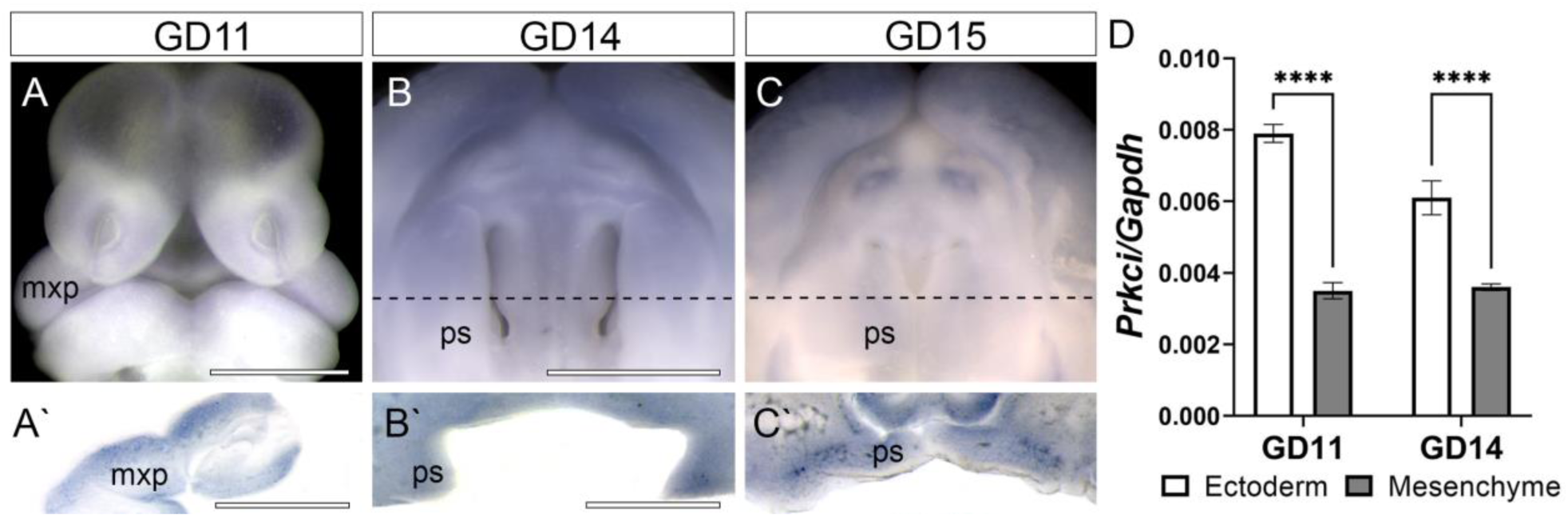
In situ hybridization shows *Prkci* expression in the mesenchyme and ectoderm during orofacial morphogenesis. A) Whole mount tissue of the face and A’) coronal section of the maxillary process (mxp) at gestational day 11 (GD11). B) Whole mount tissue B’) coronal section of the oronasal cavity and palatal shelves (ps) at GD14. C) Whole mount tissue and C’) coronal section of the palatal shelves (ps) at GD15. D) *Prkci* expression by for separated mesenchyme and ectoderm from maxillary process/palatal shelf tissue at the indicated time points. Bar graph shows the mean ± SEM of n = 3 samples per time point. Expression in the ectoderm was significantly higher than in the mesenchyme at both time points (p<0.001). Scale bars, 1mm.

### Multiple variants from affected individuals are loss-of-function, including recurrent variant Asn383Ser

Atypical PKCs are recruited to the cell membrane where they are activated by Rho-GTPase Cdc42^71–73^. Adding a prenylation domain (CAAX) targets aPKC to the plasma membrane and converts it to a constitutively active form (aPKC-CAAX)^35,74^. Injecting *Xenopus* embryos with *aPKC-CAAX* mRNA results in an expansion of the superficial cell layer of epidermal ectoderm at the expense of more basal layers^35^**, recapitulating the role of membrane-bound aPKC in specifying the superficial epidermal ectoderm fate**^36^**. This ectodermal layer is analogous to the** *Danio* enveloping layer (EVL), which we used as a model for the periderm.

First, we found aPKC activity is necessary to induce the EVL, as has been reported for other peridermopathy-associated genes *irf6*^24^ and *poky*/*chuk*/*ikk1*^33^. We also found that injecting *lacZ* mRNA or mRNA encoding the reference variant of human PRKCI, i.e., *PRKCI*, had no effect on zebrafish embryos **(Fig. 4A,B)** but that injecting *PRKCI-CAAX* mRNA led to ectopic periderm (**Fig. 4C,D**), similar to injection of *grhl3* mRNA^75^. We quantified ectopic periderm by measuring *krt4* mRNA levels relative to in control-injected embryos (**Fig. 4E**). If overexpression of an allele induced ectopic EVL, it was classified as functional; if it failed to induce ectopic EVL, it was classified as a loss of function (LOF) allele and thus considered pathogenic. Utilizing this overexpression assay, we tested 12 variants for functionality, selected using a combination of *in silico* pathogenicity predictors, minor allele frequencies, inheritance patterns, and phenotype information.

**Figure 4:**
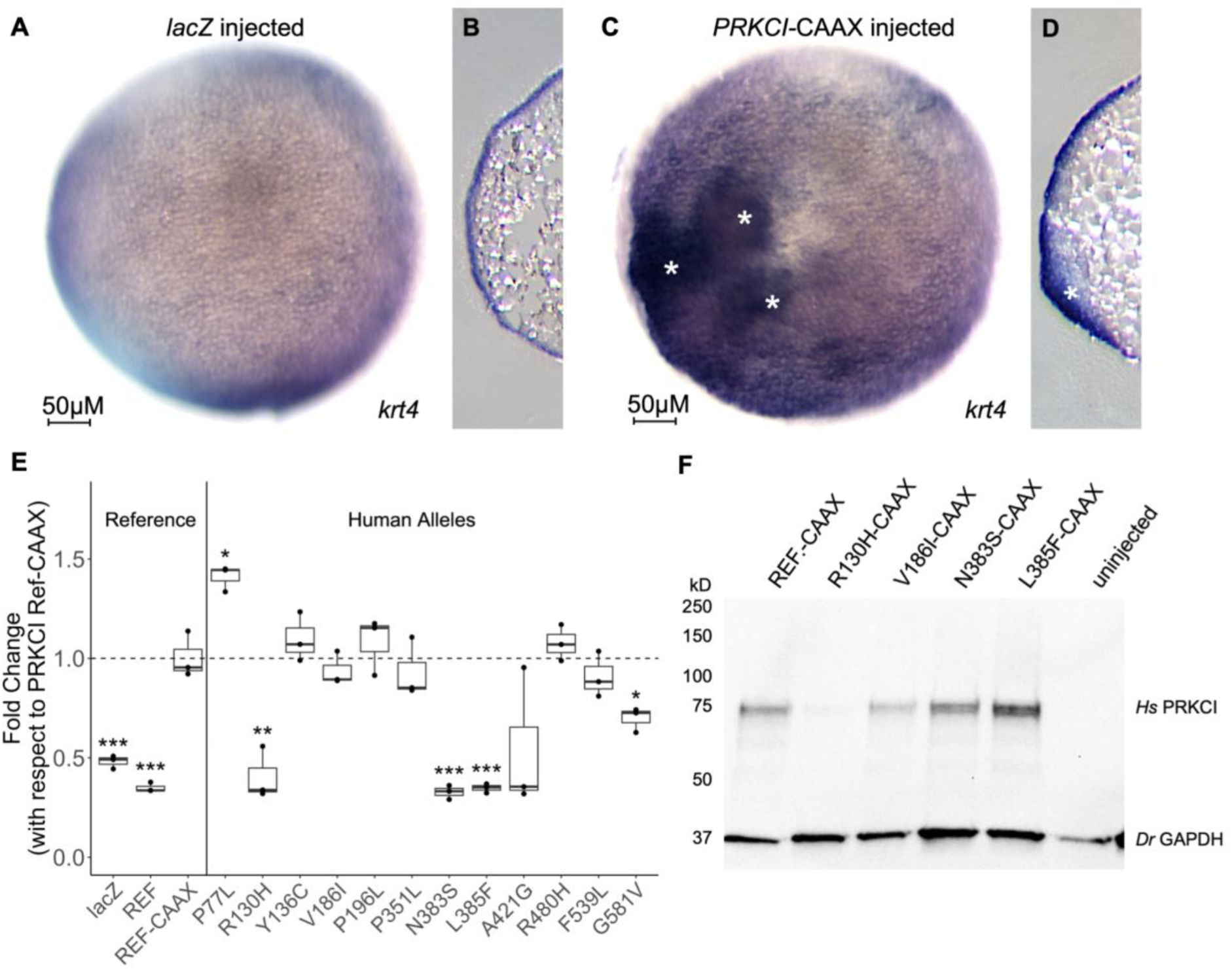
Patient-specific PRKCI alleles fail to induce ectopic enveloping layer in zebrafish embryos. A, B) LacZ injected embryo with typical enveloping layer (EVL) development at 50% epiboly. C, D) Reference PRKCI-CAAX injected embryo with ectopic EVL as indicated by white asterisks. E) Expression levels of krt4 for patient-specific alleles quantified by RT-qPCR and with respect to expression induced by reference PRCKI-CAAX. EVL is denoted by *krt4* in situ hybridization. F) Western blot showing the presence of protein in control and loss of function variants in injected embryos. *p<0.05, **p<0.005, ***p<0.001.

We injected 1-cell stage zebrafish embryos with mRNAs encoding constitutively active forms of the human PKCι reference allele and the following variants: all DNs (p.[Tyr136Cys], p.[Asn383Ser], p.[Leu385Phe]), 6 rare variants from individuals with OFCs (p.[Arg130His], p.[Pro196Leu], p.[Pro351Leu], p.[Ala421Gly], p.[Arg480His], p.[Gly581Va]l), 2 rare variants from affected individuals without an OFC (p.[Pro77Leu] and p.[Phe539Leu]), and a control variant we predicted would be benign as it was only found in an unaffected parent (p.[Val186Ile). At 50% epiboly, we measured the expression levels of *krt4*, an EVL marker, to quantify the level of ectopic expression and compared the fold change of each variant with respect to the reference mRNA to determine significant differences (**Figure 4E**).

The reference PKCι and control variant (p.[Val186Ile]) induced ectopic EVL, indicating these variants retained function, as expected. Variants p.(Pro196Leu), p.(Tyr136Cys), p.(Pro351Leu), p.(Arg480His), and p.(Phe539Leu) also induced ectopic EVL. Both p.(Ala421Gly) and p.(Gly581Val) were deemed functional although they induced less periderm than the reference *PRKCI* sequence, suggesting they may be hypomorphic. However, neither was statistically significantly different from the reference sequence. Three variants, p.(Arg130His), p.(Asn383Ser), and p.(Leu385Phe), failed to induce ectopic periderm, indicating these variants are LOF. Lastly, p.(Pro77Leu) was the only variant to induce more ectopic periderm than the reference sequence, indicating a possible gain-of-function effect.

To confirm the observed effects were due to changes in protein function, we performed Western blots for the reference and four variants: p.(Arg130His), p.(Val186Ile), p.(Asn383Ser), and p.(Leu385Phe). We injected 2-4 cell embryos with equal masses of mRNAs (separately) encoding constitutively active forms of the reference variant, harvested protein at 6 hpf, and evaluated it by Western blot. Embryos injected with each PRKCI variant had comparable levels of PRKCI protein, except those injected with p.(Arg130His), in which it was barely detectable (**Figure 4F**). However, p.(Arg130His) has been predicted to diminish PKCι stability^63^, which our results further support.

Lastly, to confirm the effects of the p.(Asn383Ser) variant, we performed an additional assay comparing the abilities of the reference variant and p.(Asn383Ser) to reverse the effects of an aPKC inhibitor. This inhibitor disrupts the EVL, leading to embryonic rupture at about 6 hpf, similar to *irf6* mutants^25^ (**Figure 5A,B**). Over-expression of PRKCI-CAAX rescued the rupture phenotype in PRKC- inhibitor treated embryos (**Figure 5C**) but Asn383Ser-CAAX lacked this ability (**Figure 5F**). These results confirm our findings in the overexpression assay, illustrating the lack of functionality of the p.(Asn383Ser) variant.

**Figure 5:**
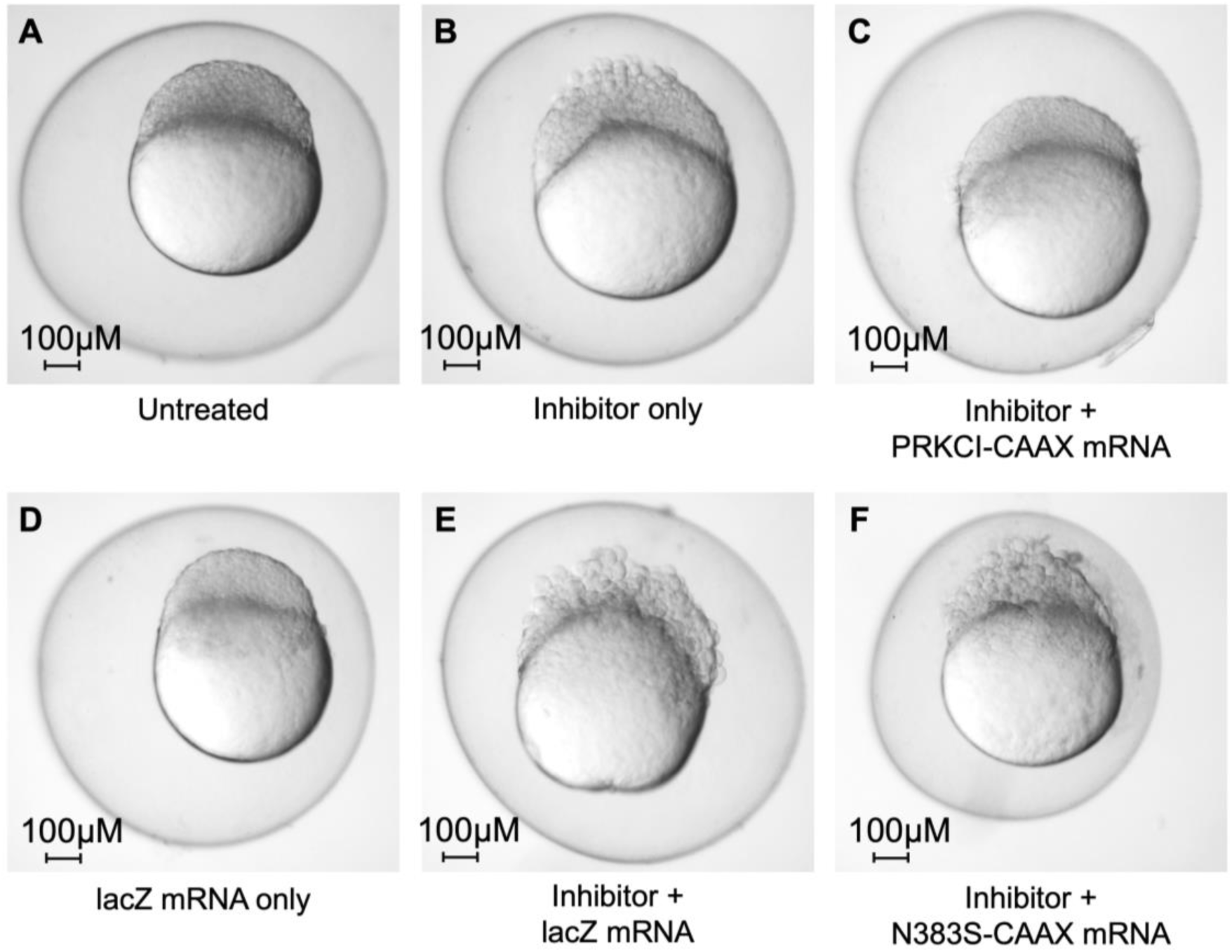
Recurrent PRKCI DN Asn383Ser fails to rescue aPKC inihibitor in zebrafish embryos treated with aPKC inhibitor. A) Control embryo with no mRNA or aPKC inhibitor. B) Embryo treated with no mRNA and aPKC inhibitor. C) Embryo treated with both aPKC inhibitor and human reference PRKCI-CAAX mRNA. D) Embryo treated with lacZ mRNA and no aPKC inhibitor. E) Embryo treated with lacZ mRNA and aPKC inhibitor. F) Embryo treated with PRKCI-N383S mRNA and aPKC inhibitor.

In summary, variants p.(Arg130His), p.(Asn383Ser), and p.(Leu385Phe) lack the ability of the PRKCI reference variant to induce ectopic EVL, which is consistent with their causing a spectrum of peridermopathy phenotypes in humans, with p.(Asn383Ser) appearing to be a variation hotspot for OFCs.. Additional characterization of the effects of the suspected gain-of-function p.(Pro77Leu) and potential hypomorphs p.(Ala421Gly) and p.(Gly581Val) are needed. Overall, our findings suggest that *PRKCI* functions within the same pathway as well-established VWS-associated genes *IRF6* and *GRHL3*, and that its disruption leads to peridermopathy and OFCs, among other phenotypes.

### Structural modelling of LOF DNs

We performed variant structure modeling for loss-of-function alleles p.(Asn383Ser) and p.(Leu385Phe) to investigate potential mechanisms of pathogenicity. Because the available crystal structure in the Protein Data Bank^59,76^ only consists of the catalytic domain, we could not model the effects of p.(Arg130His). Both Leu385 and Asn383 are proximal to the activation loop and are surface accessible (**Figure 6A**), suggesting that alteration to these residues could influence interaction with the activation loop or protein-protein interactions with PRKCI. As shown in **Figure 6B**, p.(Asn383Ser) introduces both a repulsive force ∼5A from Asp396 in the activation loop and an attractive force 4A from Lys380, potentially modifying the protein structure or disrupting binding to the activation loop. The p.(Leu385Phe) variant (**Figure 6C**) introduces an aromatic sidechain 3A from Thr395, suggesting the potential for molecular interactions. This variant also leads to introduction of a residue with a larger and bulkier sidechain near the activation loop and proximal to Gly338 and Asp339, which has the potential for disruption of the activation loop structure and surrounding residues. Given both residues are exposed, and the identified variants affect local interactions, this may provide an avenue of investigation for understanding the mechanism by which these variants result in a phenotype.

**Figure 6:**
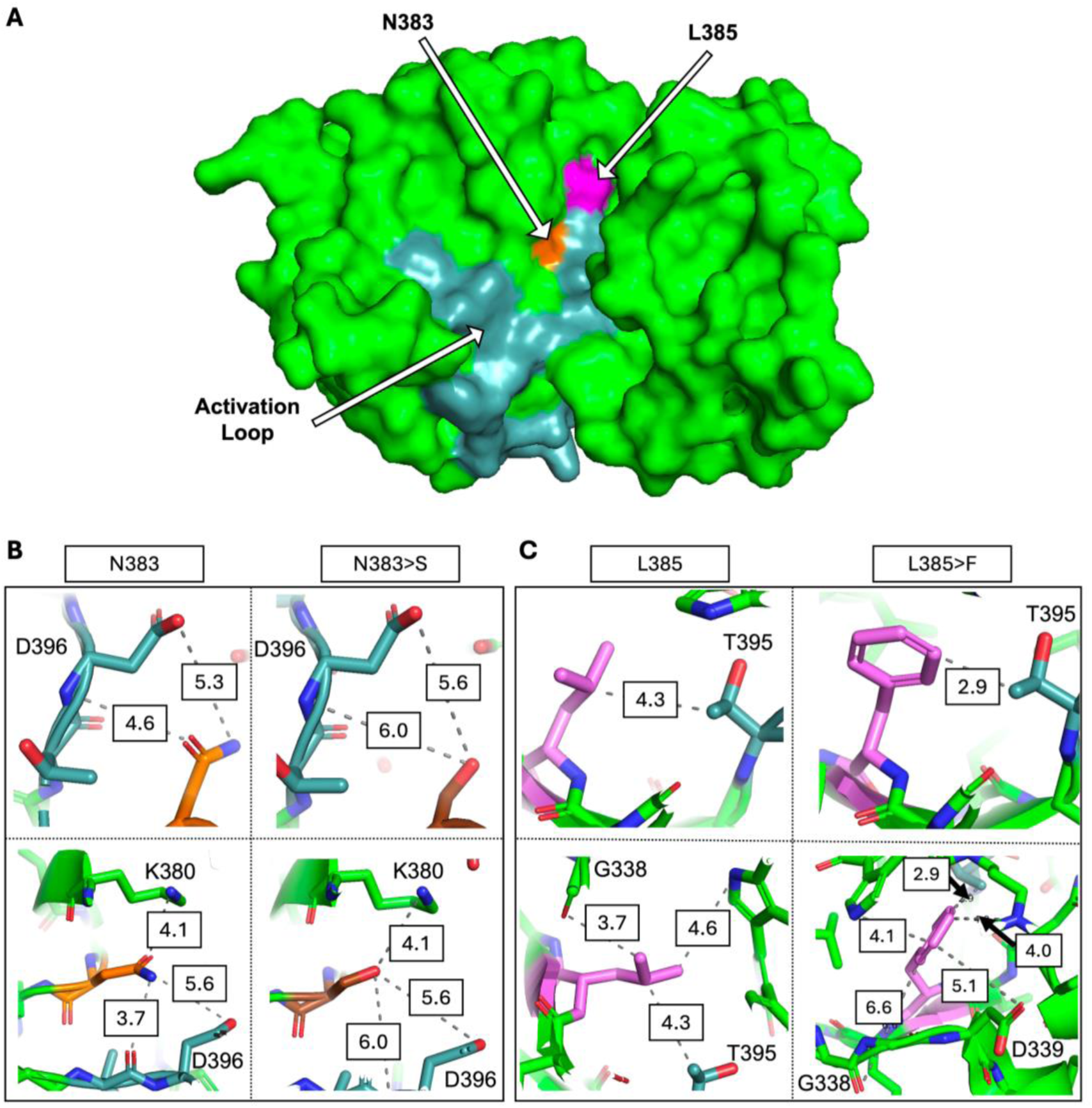
Loss-of-function variants p.(Asn383Ser) and L385 impact normal PRKCI protein structure. A) Catalytic domain rendering highlighting the activation loop (light blue) and surface-exposed variants N383 (orange) and L385 (pink). B) Variation from N383>S and C) L385>F result in altered interactions of nearby residues.

## Discussion

Here we show that variants in *PRKCI* are associated with syndromic OFCs, including VWS and other features of peridermopathy. We identified 3 variants with LOF effects in a zebrafish model, including recurrent DN p.(Asn383Ser) found in 4 unrelated individuals with OFCs. Although our initial investigation was focused on families with a clinical diagnosis of VWS and no molecular diagnosis, individuals with LOF variants exhibited a wide range of phenotypes. These include clinical features within the spectrum of peridermopathy as well as those unexplained by disruption of the periderm such as congenital heart disease, intellectual disabilities, neurodevelopmental disorders such as autism spectrum disorder, and metabolic disease.

Given the phenotypic spectrum across which peridermopathies can manifest, it is not surprising that we observed something similar in this cohort. For example, VWS and PPS are allelic disorders caused by variants in *IRF6*, sometimes the same variant, even though PPS typically manifests with more severe phenotypes^77^. Similar genetic and phenotypic overlap has been observed for other genes in the periderm TRN, including *RIPK4* implicated in both PPS^14^ and Bartsocas-Papas syndrome (BPS)^15,16^ and *CHUK* in BPS^14^ and Cocoon syndrome^17^. The range of phenotypes associated with LOF variants in *PRKCI* is striking as one individual had an isolated OFC, two had classic features of VWS, and a third had features more reminiscent of PPS. Additional individuals with variants in this gene will be needed to establish genotype-phenotype correlations, if they exist.

We also identified multiple individuals with non-peridermopathy phenotypes. Out of the individuals with rare, protein-altering variants and recorded phenotypes, 12 had intellectual and developmental disabilities (IDDs), only 1 of which co-occurred with an OFC (p.[Asn383Ser]). It is important to note, however, that our OFC cohort is varied in age of ascertainment. Although most are presumed to have isolated OFCs, individuals with OFCs are often recruited at a very young age for research studies and IDDs may become apparent at a later timepoint, resulting in misclassification of a syndromic case. Nevertheless, two of our functionally tested alleles were found in people with IDDs and no OFC (p.[Pro77Leu] and p.[Phe539Leu]). Interestingly, p.(Pro77Leu) induced significantly more ectopic EVL than the reference *PRKCI*, suggesting a gain-of-function effect. This variant is located within the PB1 domain, and although there are too few samples to establish strong conclusions, the five observed variants in this domain were all in individuals with IDDs and no OFCs. The impact of variants in this region on risk for IDDs remains unclear—our experimental model is specific to the periderm, and the effects of variation on PRKCI function in other tissues remain an area for future research. Our *in situ* staining for *PRKCI* in mouse embryos confirms expression in the palatal mesenchyme, and previous work describes PRKCI’s importance in neuronal differentiation and function^78^, so it would be reasonable to speculate that its disruption could result in the additional phenotypes.

With evidence for PKCs acting upstream of *IRF6* and *GRHL3*^28^, and aPKC being necessary for both structural analogs of the periderm in zebrafish and frog^35,36^, we hypothesize that *PRKCI* exerts its effects near the top of the known TRN. RIPK4 phosphorylates both CHUK^30^ and IRF6^28^. CHUK, in combination with IKKb, goes on to activate NF-kB^31^ while, in parallel, IRF6 activates GRHL3^26^. Further, RIPK4 is a known modulator of the PKC pathway^79^, and although current data largely describes interactions with typical PKCs, there is some evidence that RIPK4 interacts with PRKCI *in vitro*^80^. Thus, we suspect that *PRKCI* plays a role upstream of *RIPK4,* and its dysregulation may lead to decreased pathway activation. Alternatively, there is evidence that aPKC binds the IKKs to modulate NF-kB signaling^81^ to promote pro-survival signals. In its absence, there may be an increase in cell death, leading to disruption of the periderm and the observed clinical features. This discovery highlights the utility of exploring TRNs for candidate disease genes, particularly for those that have a known mechanism yet are missing molecular diagnosis in some individuals.

Our functional testing highlighted the limitations of inferring pathogenicity from in silico tools or allele frequencies. Several variants, including p.(Tyr136Cys), p.(Pro351Leu), p.(Arg480His), and p.(Phe539Leu), retained function despite predictions of pathogenicity. For p.(Tyr136Cys) and p.(Arg480His), published research provides context in which to interpret the results of the zebrafish overexpression assay. For p.(Tyr136Cys), *in vitro* evidence suggests this tyrosine plays an important role in the PKCι regulatory module and membrane binding. Previously, substituting Tyr136 for glutamic acid led to a reduced level of the PKCι kinase domain in HEK293T cells, presumably due to a decreased thermostability of the protein^62^. Because our model includes a membrane-guided tag, the zebrafish overexpression assay may be unable to capture effects secondary to defective membrane binding, and the effect of p.(Tyr136Cys) should be considered inconclusive for this assay.

The substitution p.Arg480His has also been reported as a somatic mutation in some cancers, though p.(Arg480Cys) is more common in the Catalog of Somatic Mutations in Cancer (COSMIC, cancer.sanger.ac.uk)^82^. This change (p.Arg480Cys) is reported to result in a loss of substrate specificity^65^, and it is possible that a similar effect could occur in our observed p.(Arg480His) variant. The individuals’ phenotypes included one with isolated CP and another with severe intrauterine growth restriction and multiple congenital heart defects. The result of our assay may indicate that this variant is not pathogenic, though it is also possible that p.(Arg480His) results in effects in tissues outside the periderm which we are unable to detect in this model.

We also identified two variants as possible hypomorphs, p.(Ala421Gly) and p.(Gly581Val), though it is important to note that our functional testing method is sensitive to variants with full loss-of-function effects and does not easily allow for measurement of more nuanced effects. While there is little in the literature to aid interpretation for p.(Ala421Gly), it is novel in gnomAD 4.1.0 and predicted to be deleterious by SIFT, PolyPhen2, and AlphaMissense. Similarly, p.(Gly581Val) is rare in gnomAD (MAF 1.97x10^-5^) and is predicted deleterious by the same algorithms but was also among the most likely missense variants to be deleterious on the basis of *in silico* predictions of effects on protein stability and conformational dynamics^63^. Although we cannot draw strong conclusions about p.(Ala421Gly) and p.Gly581Val, both variants warrant additional study to determine their pathogenicity. Ideally, a second model system would help further elucidate functionality of these variants, however, due to the early lethality in murine models, this may be limited to *in vitro* modeling or using conditional approaches.

Interestingly, p.(Arg130His) was found in a family with an affected proband and mother, yet it was inherited from the unaffected father. Based on this inheritance pattern, we suspected this variant would retain function and found the opposite to be true. It is worth noting that p.(Arg130His) has been predicted to diminish PKCι stability^63^, which may indicate a more complex genetic background in this specific individual. In contrast, p.(Asn383Ser) appears to be a mutation hotspot, although it is unclear what is driving the recurrence of this variant. Common mechanisms that contribute to mutation hotspots are GC-rich and highly repetitive sequences^83^, but neither applies at this residue; thus, the common mechanisms of alteration^83^ are unlikely factors at this residue.

Aside from zebrafish, mice are often used as a model organism for OFCs. However, *PRKCI* plays vital roles in early development, which makes using a murine model challenging as knockout results in early embryonic lethality. Loss of *Prkci* in embryoid bodies results in failed cavitation^84^ and homozygous knockout of *Prkci* (which encodes PKCλ in mice) leads to embryonic lethality by day E9.5 due to polarity defects^85^. A second challenge is that while homozygous knockouts are essentially lethal at full penetrance, heterozygous mice do not exhibit abnormal phenotypes. However, all our variants of interest are heterozygous in affected individuals. While this is consistent with an autosomal dominant phenotype like VWS, it precludes mice as an easy model for study.

When we compare these findings to human data, such as the genome aggregate database (gnomAD) v4.1.0, we find moderate intolerance to loss-of-function with a LOEUF score of 0.66 (threshold of <0.6 suggested for interpreting Mendelian diseases) and a missense Z-score of 3.07, which is just below the threshold of 3.09 for significant constraint. For comparison, *IRF6* and *GRHL3* have LOEUF scores of 0.21 and 0.4, and missense Z-scores of 3.9 and 1.72, respectively. Although we expected a lower LOEUF score based on mouse lethality, there is still a depletion of both LOF and missense variants cumulatively across *PRKCI*. It is possible that disruption of a specific domain is more lethal, for example loss of PB1 leading to cell polarity defects; however, there is currently no significant regional constraint in gnomAD to support this hypothesis. Alternatively, there may be unknown redundant functions between aPKC genes in humans, as has been shown for aPKC zeta (PRKζ) and PRKCI in mice^85,86^. Lastly, there may be additional modifiers not yet identified that contribute to both penetrance and expressivity of observed phenotypes in individuals harboring *PRKCI* variants. Regardless, on an individual basis, all our confirmed LOF alleles are rare or novel, heterozygous, missense variants with evidence for causality.

Taken altogether, we provide evidence that variants in *PRKCI* are causal for VWS and other features of peridermopathies, with p.(Asn383Ser) being a particular hotspot. Of the ∼20% of VWS cases without a genetic diagnosis, we estimate around 2-3% may be due to variants in *PRKCI*. Likewise, as *PRKCI* appears to work within the same periderm network as previously associated genes *IRF6* and *GRHL3*, we suspect additional candidate genes can be found within this TRN to further explain undiagnosed cases. Ultimately our findings contribute a novel gene to VWS associations and provide an avenue of future investigation for both additional VWS cases and the role of PRKCI in other phenotypes.

## Data Availability

Sequence and phenotype data is available from the Database of Genotypes and Phenotypes (dbGaP, ncbi.nlm.nih.gov) under study accession # phs002220.v1.p1 (CPSeq, VWS probands), accession # phs001420.v1.p1 (Latin American), accession # phs001168.v2.p2 (US, European), accession # phs001997,v1.p1 (African, Asian). Additional information for probands in the GMKF cohort can be viewed at https://kidsfirstdrc.org/studies/.

## Acknowledgements

We are extremely grateful to the participants and their families as well as colleagues who have made this research possible. Sequencing services for CPSeq were provided by the Center for Inherited Disease Research (CIDR). CIDR is fully funded through a federal contract from the National Institutes of Health to The Johns Hopkins University, contract number HHSN268201700006I. Additional whole genome sequencing was funded by National Institutes of Health (NIH) grants: X01-HL136465 (MLM: US, European trios), X01-HL132363 (MLM: Colombian trios), X01-DE0300701 (MLM: Latin American trios), and X01-HG010835 (EJL, MLM: CPseq trios). Patient recruitment, assembly of phenotypic information, and data analysis were supported by National Institutes of Health (NIH) grants: F31-DE032588 (KR), X01-HG010835 (EJL), R01 DE027983 (EJL), R01 DE028342 (EJL), R01 DE030342 (EJL), R01-DE028300 (AB), R01DE032710 (RJL), DE008559 (JCM), R01-DE016148 (MLM, SMW), R01-DE008559 (JCM, MLM), R01-DE032122 (MLM), R01-DE0332319 (MLM).

## Author contributions

This study conceptualized by EJL, RAC, and RJL. Resources were contributed by WLA, THB, AB, CJB, WKC, BG, LJJG, JTH, LMU, JCM, DAS, GMS, MAT, SMW, HB, MLM, RJL, RAC, and EJL. Data curation was performed by KR, SKS, RBW, DJC, HB, RJL, and RAC. Data analysis, investigation, and visualization were performed by KR, SKS, RBW, DVF, AF under supervision of DJC, MPE, RJL, RAC, EJL. The originally manuscript was primarily drafted by KR with additional contributions from SKS, RBW, DVF, and AF. All authors contributed to critical review and approval of the final draft.

## References

1. Dronamraju, K.R. (1971). Genetic studies of a cleft palate clinic population. Birth Defects Orig Artic Ser 7, 54–57.

2. Murray, J.C., Daack-Hirsch, S., Buetow, K.H., Munger, R., Espina, L., Paglinawan, N., Villanueva, E., Rary, J., Magee, K., and Magee, W. (1997). Clinical and epidemiologic studies of cleft lip and palate in the Philippines. Cleft Palate Craniofac J 34, 7–10. 10.1597/1545-1569_1997_034_0007_caesoc_2.3.co_2.

3. Van Der Woude, A. (1954). Fistula labii inferioris congenita and its association with cleft lip and palate. Am J Hum Genet 6, 244–256.

4. Janku, P., Robinow, M., Kelly, T., Bralley, R., Baynes, A., and Edgerton, M.T. (1980). The van der Woude syndrome in a large kindred: variability, penetrance, genetic risks. Am J Med Genet 5, 117–123. 10.1002/ajmg.1320050203.

5. Tan, E.C., Lim, E.C., and Lee, S.T. (2013). De novo 2.3 Mb microdeletion of 1q32.2 involving the Van der Woude Syndrome locus. Mol Cytogenet 6, 31. 10.1186/1755-8166-6-31.

6. Sander, A., Schmelzle, R., and Murray, J. (1994). Evidence for a microdeletion in 1q32-41 involving the gene responsible for Van der Woude syndrome. Hum Mol Genet 3, 575–578. 10.1093/hmg/3.4.575.

7. Al-Kurbi, A.A., Aliyev, E., AlSa’afin, S., Aamer, W., Palaniswamy, S., Al-Maraghi, A., Kilani, H., Akil, A.A., Stotland, M.A., and Fakhro, K.A. (2023). A Complex Intrachromosomal Rearrangement Disrupting IRF6 in a Family with Popliteal Pterygium and Van der Woude Syndromes. Genes (Basel) 14. 10.3390/genes14040849.

8. Kondo, S., Schutte, B.C., Richardson, R.J., Bjork, B.C., Knight, A.S., Watanabe, Y., Howard, E., de Lima, R.L., Daack-Hirsch, S., Sander, A., et al. (2002). Mutations in IRF6 cause Van der Woude and popliteal pterygium syndromes. Nat Genet 32, 285–289. 10.1038/ng985.

9. de Lima, R.L., Hoper, S.A., Ghassibe, M., Cooper, M.E., Rorick, N.K., Kondo, S., Katz, L., Marazita, M.L., Compton, J., Bale, S., et al. (2009). Prevalence and nonrandom distribution of exonic mutations in interferon regulatory factor 6 in 307 families with Van der Woude syndrome and 37 families with popliteal pterygium syndrome. Genet Med 11, 241–247. 10.1097/GIM.0b013e318197a49a.

10. Leslie, E.J., Standley, J., Compton, J., Bale, S., Schutte, B.C., and Murray, J.C. (2013). Comparative analysis of IRF6 variants in families with Van der Woude syndrome and popliteal pterygium syndrome using public whole-exome databases. Genet Med 15, 338–344. 10.1038/gim.2012.141.

11. Peyrard-Janvid, M., Leslie, E.J., Kousa, Y.A., Smith, T.L., Dunnwald, M., Magnusson, M., Lentz, B.A., Unneberg, P., Fransson, I., Koillinen, H.K., et al. (2014). Dominant mutations in GRHL3 cause Van der Woude Syndrome and disrupt oral periderm development. Am J Hum Genet 94, 23–32. 10.1016/j.ajhg.2013.11.009.

12. Richardson, R.J., Hammond, N.L., Coulombe, P.A., Saloranta, C., Nousiainen, H.O., Salonen, R., Berry, A., Hanley, N., Headon, D., Karikoski, R., and Dixon, M.J. (2014). Periderm prevents pathological epithelial adhesions during embryogenesis. J Clin Invest 124, 3891–3900. 10.1172/jci71946.

13. Froster-Iskenius, U.G. (1990). Popliteal pterygium syndrome. J Med Genet 27, 320–326. 10.1136/jmg.27.5.320.

14. Leslie, E.J., O’Sullivan, J., Cunningham, M.L., Singh, A., Goudy, S.L., Ababneh, F., Alsubaie, L., Ch’ng, G.S., van der Laar, I.M., Hoogeboom, A.J., et al. (2015). Expanding the genetic and phenotypic spectrum of popliteal pterygium disorders. Am J Med Genet A *167A*, 545-552. 10.1002/ajmg.a.36896.

15. Kalay, E., Sezgin, O., Chellappa, V., Mutlu, M., Morsy, H., Kayserili, H., Kreiger, E., Cansu, A., Toraman, B., Abdalla, E.M., et al. (2012). Mutations in RIPK4 cause the autosomal-recessive form of popliteal pterygium syndrome. Am J Hum Genet 90, 76–85. 10.1016/j.ajhg.2011.11.014.

16. Mitchell, K., O’Sullivan, J., Missero, C., Blair, E., Richardson, R., Anderson, B., Antonini, D., Murray, J.C., Shanske, A.L., Schutte, B.C., et al. (2012). Exome sequence identifies RIPK4 as the Bartsocas-Papas syndrome locus. Am J Hum Genet 90, 69–75. 10.1016/j.ajhg.2011.11.013.

17. Lahtela, J., Nousiainen, H.O., Stefanovic, V., Tallila, J., Viskari, H., Karikoski, R., Gentile, M., Saloranta, C., Varilo, T., Salonen, R., and Kestilä, M. (2010). Mutant CHUK and Severe Fetal Encasement Malformation. New Engl J Med 363, 1631–1637. 10.1056/NEJMoa0911698.

18. Kimmel, C.B., Warga, R.M., and Schilling, T.F. (1990). Origin and organization of the zebrafish fate map. Development 108, 581–594. 10.1242/dev.108.4.581.

19. De Groote, P., Tran, H.T., Fransen, M., Tanghe, G., Urwyler, C., De Craene, B., Leurs, K., Gilbert, B., Van Imschoot, G., De Rycke, R., et al. (2015). A novel RIPK4-IRF6 connection is required to prevent epithelial fusions characteristic for popliteal pterygium syndromes. Cell Death Differ 22, 1012–1024. 10.1038/cdd.2014.191.

20. Carroll, S.H., Schafer, S., Dalessandro, E., Ho, T.V., Chai, Y., and Liao, E.C. (2024). Neural crest and periderm-specific requirements of Irf6 during neural tube and craniofacial development. bioRxiv. 10.1101/2024.06.11.598425.

21. Ingraham, C.R., Kinoshita, A., Kondo, S., Yang, B., Sajan, S., Trout, K.J., Malik, M.I., Dunnwald, M., Goudy, S.L., Lovett, M., et al. (2006). Abnormal skin, limb and craniofacial morphogenesis in mice deficient for interferon regulatory factor 6 (Irf6). Nat Genet 38, 1335–1340. 10.1038/ng1903.

22. Richardson, R.J., Dixon, J., Malhotra, S., Hardman, M.J., Knowles, L., Boot-Handford, R.P., Shore, P., Whitmarsh, A., and Dixon, M.J. (2006). Irf6 is a key determinant of the keratinocyte proliferation-differentiation switch. Nat Genet 38, 1329–1334. 10.1038/ng1894.

23. Kashgari, G., Meinecke, L., Gordon, W., Ruiz, B., Yang, J., Ma, A.L., Xie, Y., Ho, H., Plikus, M.V., Nie, Q., et al. (2020). Epithelial Migration and Non-adhesive Periderm Are Required for Digit Separation during Mammalian Development. Dev Cell 52, 764–778.e764. 10.1016/j.devcel.2020.01.032.

24. Sabel, J.L., d’Alençon, C., O’Brien, E.K., Van Otterloo, E., Lutz, K., Cuykendall, T.N., Schutte, B.C., Houston, D.W., and Cornell, R.A. (2009). Maternal Interferon Regulatory Factor 6 is required for the differentiation of primary superficial epithelia in Danio and Xenopus embryos. Developmental Biology *325*, 249-262. 10.1016/j.ydbio.2008.10.031.

25. Li, E.B., Truong, D., Hallett, S.A., Mukherjee, K., Schutte, B.C., and Liao, E.C. (2017). Rapid functional analysis of computationally complex rare human IRF6 gene variants using a novel zebrafish model. PLoS Genet 13, e1007009. 10.1371/journal.pgen.1007009.

26. de la Garza, G., Schleiffarth, J.R., Dunnwald, M., Mankad, A., Weirather, J.L., Bonde, G., Butcher, S., Mansour, T.A., Kousa, Y.A., Fukazawa, C.F., et al. (2013). Interferon regulatory factor 6 promotes differentiation of the periderm by activating expression of Grainyhead-like 3. J Invest Dermatol 133, 68–77. 10.1038/jid.2012.269.

27. Miles, L.B., Darido, C., Kaslin, J., Heath, J.K., Jane, S.M., and Dworkin, S. (2017). Mis-expression of grainyhead-like transcription factors in zebrafish leads to defects in enveloping layer (EVL) integrity, cellular morphogenesis and axial extension. Sci Rep 7, 17607. 10.1038/s41598-017-17898-7.

28. Kwa, M.Q., Huynh, J., Aw, J., Zhang, L., Nguyen, T., Reynolds, E.C., Sweet, M.J., Hamilton, J.A., and Scholz, G.M. (2014). Receptor-interacting protein kinase 4 and interferon regulatory factor 6 function as a signaling axis to regulate keratinocyte differentiation. J Biol Chem 289, 31077–31087. 10.1074/jbc.M114.589382.

29. Oberbeck, N., Pham, V.C., Webster, J.D., Reja, R., Huang, C.S., Zhang, Y., Roose-Girma, M., Warming, S., Li, Q., Birnberg, A., et al. (2019). The RIPK4-IRF6 signalling axis safeguards epidermal differentiation and barrier function. Nature 574, 249–253. 10.1038/s41586-019-1615-3.

30. Kim, S.W., Schifano, M., Oleksyn, D., Jordan, C.T., Ryan, D., Insel, R., Zhao, J., and Chen, L. (2014). Protein kinase C-associated kinase regulates NF-κB activation through inducing IKK activation. Int J Oncol 45, 1707–1714. 10.3892/ijo.2014.2578.

31. Seitz, C.S., Freiberg, R.A., Hinata, K., and Khavari, P.A. (2000). NF-kappaB determines localization and features of cell death in epidermis. J Clin Invest 105, 253–260. 10.1172/jci7630.

32. Hu, Y., Baud, V., Delhase, M., Zhang, P., Deerinck, T., Ellisman, M., Johnson, R., and Karin, M. (1999). Abnormal morphogenesis but intact IKK activation in mice lacking the IKKalpha subunit of IkappaB kinase. Science 284, 316–320. 10.1126/science.284.5412.316.

33. Fukazawa, C., Santiago, C., Park, K.M., Deery, W.J., Gomez de la Torre Canny, S., Holterhoff, C.K., and Wagner, D.S. (2010). poky/chuk/ikk1 is required for differentiation of the zebrafish embryonic epidermis. Developmental Biology 346, 272–283. 10.1016/j.ydbio.2010.07.037.

34. Tanghe, G., Urwyler-Rösselet, C., De Groote, P., Dejardin, E., De Bock, P.J., Gevaert, K., Vandenabeele, P., and Declercq, W. (2018). RIPK4 activity in keratinocytes is controlled by the SCF(β-TrCP) ubiquitin ligase to maintain cortical actin organization. Cell Mol Life Sci 75, 2827–2841. 10.1007/s00018-018-2763-6.

35. Ossipova, O., Tabler, J., Green, J.B., and Sokol, S.Y. (2007). PAR1 specifies ciliated cells in vertebrate ectoderm downstream of aPKC. Development 134, 4297–4306. 10.1242/dev.009282.

36. Chalmers, A.D., Strauss, B., and Papalopulu, N. (2003). Oriented cell divisions asymmetrically segregate aPKC and generate cell fate diversity in the early Xenopus embryo. Development 130, 2657–2668. 10.1242/dev.00490.

37. Robinson, K., Mosley, T.J., Rivera-González, K.S., Jabbarpour, C.R., Curtis, S.W., Adeyemo, W.L., Beaty, T.H., Butali, A., Buxó, C.J., Cutler, D.J., et al. (2023). Trio-based GWAS identifies novel associations and subtype-specific risk factors for cleft palate. Human Genetics and Genomics Advances 4. 10.1016/j.xhgg.2023.100234.

38. Bishop, M.R., Diaz Perez, K.K., Sun, M., Ho, S., Chopra, P., Mukhopadhyay, N., Hetmanski, J.B., Taub, M.A., Moreno-Uribe, L.M., Valencia-Ramirez, L.C., et al. (2020). Genome-wide Enrichment of De Novo Coding Mutations in Orofacial Cleft Trios. Am J Hum Genet 107, 124–136. 10.1016/j.ajhg.2020.05.018.

39. Conrad, D.F., Keebler, J.E., DePristo, M.A., Lindsay, S.J., Zhang, Y., Casals, F., Idaghdour, Y., Hartl, C.L., Torroja, C., Garimella, K.V., et al. (2011). Variation in genome-wide mutation rates within and between human families. Nat Genet 43, 712–714. 10.1038/ng.862.

40. McKenna, A., Hanna, M., Banks, E., Sivachenko, A., Cibulskis, K., Kernytsky, A., Garimella, K., Altshuler, D., Gabriel, S., Daly, M., and DePristo, M.A. (2010). The Genome Analysis Toolkit: a MapReduce framework for analyzing next-generation DNA sequencing data. Genome Res 20, 1297–1303. 10.1101/gr.107524.110.

41. Van der Auwera, G.A., Carneiro, M.O., Hartl, C., Poplin, R., Del Angel, G., Levy-Moonshine, A., Jordan, T., Shakir, K., Roazen, D., Thibault, J., et al. (2013). From FastQ data to high confidence variant calls: the Genome Analysis Toolkit best practices pipeline. Curr Protoc Bioinformatics 43, 11 10 11-11 10 33. 10.1002/0471250953.bi1110s43.

42. Mukhopadhyay, N., Bishop, M., Mortillo, M., Chopra, P., Hetmanski, J.B., Taub, M.A., Moreno, L.M., Valencia-Ramirez, L.C., Restrepo, C., Wehby, G.L., et al. (2020). Whole genome sequencing of orofacial cleft trios from the Gabriella Miller Kids First Pediatric Research Consortium identifies a new locus on chromosome 21. Hum Genet 139, 215–226. 10.1007/s00439-019-02099-1.

43. Wang, K., Li, M., and Hakonarson, H. (2010). ANNOVAR: functional annotation of genetic variants from high-throughput sequencing data. Nucleic Acids Res 38, e164. 10.1093/nar/gkq603.

44. Ng, P.C., and Henikoff, S. (2003). SIFT: Predicting amino acid changes that affect protein function. Nucleic Acids Res 31, 3812–3814. 10.1093/nar/gkg509.

45. Adzhubei, I.A., Schmidt, S., Peshkin, L., Ramensky, V.E., Gerasimova, A., Bork, P., Kondrashov, A.S., and Sunyaev, S.R. (2010). A method and server for predicting damaging missense mutations. Nat Methods 7, 248–249. 10.1038/nmeth0410-248.

46. Cheng, J., Novati, G., Pan, J., Bycroft, C., Žemgulytė, A., Applebaum, T., Pritzel, A., Wong, L.H., Zielinski, M., Sargeant, T., et al. (2023). Accurate proteome-wide missense variant effect prediction with AlphaMissense. Science 381, eadg7492. doi:10.1126/science.adg7492.

47. Samocha, K.E., Robinson, E.B., Sanders, S.J., Stevens, C., Sabo, A., McGrath, L.M., Kosmicki, J.A., Rehnstrom, K., Mallick, S., Kirby, A., et al. (2014). A framework for the interpretation of de novo mutation in human disease. Nat Genet 46, 944–950. 10.1038/ng.3050.

48. Kimmel, C.B., Ballard, W.W., Kimmel, S.R., Ullmann, B., and Schilling, T.F. (1995). Stages of embryonic development of the zebrafish. Dev Dyn 203, 253–310. 10.1002/aja.1002030302.

49. Cokus, S.J., De La Torre, M., Medina, E.F., Rasmussen, J.P., Ramirez-Gutierrez, J., Sagasti, A., and Wang, F. (2019). Tissue-Specific Transcriptomes Reveal Gene Expression Trajectories in Two Maturing Skin Epithelial Layers in Zebrafish Embryos. G3 Genes|Genomes|Genetics *9*, 3439-3452. 10.1534/g3.119.400402.

50. Livak, K.J., and Schmittgen, T.D. (2001). Analysis of relative gene expression data using real-time quantitative PCR and the 2(-Delta Delta C(T)) Method. Methods 25, 402–408. 10.1006/meth.2001.1262.

51. Westerfield, M. (2007). THE ZEBRAFISH BOOK, 5th Edition; A guide for the laboratory use of zebrafish (Danio rerio) (University of Oregon Press, Eugene).

52. Heyne, G.W., Plisch, E.H., Melberg, C.G., Sandgren, E.P., Peter, J.A., and Lipinski, R.J. (2015). A Simple and Reliable Method for Early Pregnancy Detection in Inbred Mice. J Am Assoc Lab Anim Sci 54, 368–371.

53. Everson, J.L., Sun, M.R., Fink, D.M., Heyne, G.W., Melberg, C.G., Nelson, K.F., Doroodchi, P., Colopy, L.J., Ulschmid, C.M., Martin, A.A., et al. (2019). Developmental Toxicity Assessment of Piperonyl Butoxide Exposure Targeting Sonic Hedgehog Signaling and Forebrain and Face Morphogenesis in the Mouse: An in Vitro and in Vivo Study. Environ Health Perspect 127, 107006. 10.1289/EHP5260.

54. Abler, L.L., Mehta, V., Keil, K.P., Joshi, P.S., Flucus, C.L., Hardin, H.A., Schmitz, C.T., and Vezina, C.M. (2011). A high throughput in situ hybridization method to characterize mRNA expression patterns in the fetal mouse lower urogenital tract. J Vis Exp. 10.3791/2912.

55. Everson, J.L., Fink, D.M., Yoon, J.W., Leslie, E.J., Kietzman, H.W., Ansen-Wilson, L.J., Chung, H.M., Walterhouse, D.O., Marazita, M.L., and Lipinski, R.J. (2017). Sonic hedgehog regulation of Foxf2 promotes cranial neural crest mesenchyme proliferation and is disrupted in cleft lip morphogenesis. Development 144, 2082–2091. 10.1242/dev.149930.

56. Li, H., and Williams, T. (2013). Separation of mouse embryonic facial ectoderm and mesenchyme. J Vis Exp. 10.3791/50248.

57. Rizos, M., and Spyropoulos, M.N. (2004). Van der Woude syndrome: a review. Cardinal signs, epidemiology, associated features, differential diagnosis, expressivity, genetic counselling and treatment. European Journal of Orthodontics 26, 17–24. 10.1093/ejo/26.1.17.

58. Sobreira, N., Schiettecatte, F., Valle, D., and Hamosh, A. (2015). GeneMatcher: a matching tool for connecting investigators with an interest in the same gene. Hum Mutat 36, 928–930. 10.1002/humu.22844.

59. Messerschmidt, A., Macieira, S., Velarde, M., Bädeker, M., Benda, C., Jestel, A., Brandstetter, H., Neuefeind, T., and Blaesse, M. (2005). Crystal Structure of the Catalytic Domain of Human Atypical Protein Kinase C-iota Reveals Interaction Mode of Phosphorylation Site in Turn Motif. Journal of Molecular Biology 352, 918–931. 10.1016/j.jmb.2005.07.060.

60. Zhou, X., Edmonson, M.N., Wilkinson, M.R., Patel, A., Wu, G., Liu, Y., Li, Y., Zhang, Z., Rusch, M.C., Parker, M., et al. (2016). Exploring genomic alteration in pediatric cancer using ProteinPaint. Nat Genet 48, 4–6. 10.1038/ng.3466.

61. Manning, G., Whyte, D.B., Martinez, R., Hunter, T., and Sudarsanam, S. (2002). The Protein Kinase Complement of the Human Genome. Science 298, 1912–1934. doi:10.1126/science.1075762.

62. Cobbaut, M., McDonald, N.Q., and Parker, P.J. (2023). Control of atypical PKCι membrane dissociation by tyrosine phosphorylation within a PB1-C1 interdomain interface. J Biol Chem 299, 104847. 10.1016/j.jbc.2023.104847.

63. Shah, H., Khan, K., Khan, N., Badshah, Y., Ashraf, N.M., and Shabbir, M. (2022). Impact of deleterious missense PRKCI variants on structural and functional dynamics of protein. Sci Rep-Uk 12, 3781. 10.1038/s41598-022-07526-4.

64. Linch, M., Sanz-Garcia, M., Soriano, E., Zhang, Y., Riou, P., Rosse, C., Cameron, A., Knowles, P., Purkiss, A., Kjaer, S., et al. (2013). A Cancer-Associated Mutation in Atypical Protein Kinase Cι Occurs in a Substrate-Specific Recruitment Motif. Science Signaling 6, ra82-ra82. doi:10.1126/scisignal.2004068.

65. Reina-Campos, M., Diaz-Meco, M.T., and Moscat, J. (2019). The Dual Roles of the Atypical Protein Kinase Cs in Cancer. Cancer Cell 36, 218–235. 10.1016/j.ccell.2019.07.010.

66. Moscat, J., Diaz-Meco, M.T., Albert, A., and Campuzano, S. (2006). Cell signaling and function organized by PB1 domain interactions. Mol Cell 23, 631–640. 10.1016/j.molcel.2006.08.002.

67. Steinberg, S.F. (2008). Structural basis of protein kinase C isoform function. Physiol Rev 88, 1341–1378. 10.1152/physrev.00034.2007.

68. Niessen, M.T., Scott, J., Zielinski, J.G., Vorhagen, S., Sotiropoulou, P.A., Blanpain, C., Leitges, M., and Niessen, C.M. (2013). aPKCλ controls epidermal homeostasis and stem cell fate through regulation of division orientation. J Cell Biol 202, 887–900. 10.1083/jcb.201307001.

69. Yankee, T.N., Oh, S., Winchester, E.W., Wilderman, A., Robinson, K., Gordon, T., Rosenfeld, J.A., VanOudenhove, J., Scott, D.A., Leslie, E.J., and Cotney, J. (2023). Integrative analysis of transcriptome dynamics during human craniofacial development identifies candidate disease genes. Nat Commun 14, 4623. 10.1038/s41467-023-40363-1.

70. Kovac, J., Oster, H., and Leitges, M. (2007). Expression of the atypical protein kinase C (aPKC) isoforms ι/λ and ζ during mouse embryogenesis. Gene Expression Patterns 7, 187–196. 10.1016/j.modgep.2006.07.002.

71. Joberty, G., Petersen, C., Gao, L., and Macara, I.G. (2000). The cell-polarity protein Par6 links Par3 and atypical protein kinase C to Cdc42. Nature Cell Biology 2, 531–539. 10.1038/35019573.

72. Lin, D., Edwards, A.S., Fawcett, J.P., Mbamalu, G., Scott, J.D., and Pawson, T. (2000). A mammalian PAR-3-PAR-6 complex implicated in Cdc42/Rac1 and aPKC signalling and cell polarity. Nat Cell Biol 2, 540–547. 10.1038/35019582.

73. Sabherwal, N., Tsutsui, A., Hodge, S., Wei, J., Chalmers, A.D., and Papalopulu, N. (2009). The apicobasal polarity kinase aPKC functions as a nuclear determinant and regulates cell proliferation and fate during Xenopus primary neurogenesis. Development 136, 2767–2777. 10.1242/dev.034454.

74. Lee, C.Y., Robinson, K.J., and Doe, C.Q. (2006). Lgl, Pins and aPKC regulate neuroblast self-renewal versus differentiation. Nature 439, 594–598. 10.1038/nature04299.

75. Eshete, M.A., Liu, H., Li, M., Adeyemo, W.L., Gowans, L.J.J., Mossey, P.A., Busch, T., Deressa, W., Donkor, P., Olaitan, P.B., et al. (2018). Loss-of-Function GRHL3 Variants Detected in African Patients with Isolated Cleft Palate. J Dent Res 97, 41–48. 10.1177/0022034517729819.

76. Messerschmidt, A., Macieira, S., Velarde, M., Baedeker, M., Benda, C., Jestel, A., Brandstetter, H., Neuefeind, T., Blaesse, M., Structural Proteomics in Europe (SPINE) (2005). Crystal Structure of the Catalytic Domain of Atypical Protein Kinase C-iota. 10.2210/pdb1ZRZ/pdb.

77. Schutte, B.C., Saal, H.M., Goudy, S., and Leslie, E.J. (1993). IRF6-Related Disorders. In GeneReviews((R)), M.P. Adam, H.H. Ardinger, R.A. Pagon, S.E. Wallace, L.J.H. Bean, G. Mirzaa, and A. Amemiya, eds.

78. Hapak, S.M., Rothlin, C.V., and Ghosh, S. (2019). aPKC in neuronal differentiation, maturation and function. Neuronal Signal 3, Ns20190019. 10.1042/ns20190019.

79. Bähr, C., Rohwer, A., Stempka, L., Rincke, G., Marks, F., and Gschwendt, M. (2000). DIK, a Novel Protein Kinase That Interacts with Protein Kinase C&#x3b4;: CLONING, CHARACTERIZATION, AND GENE ANALYSIS *. Journal of Biological Chemistry 275, 36350–36357. 10.1074/jbc.M004771200.

80. Rodriguez, J., Pilkington, R., Garcia Munoz, A., Nguyen, Lan K., Rauch, N., Kennedy, S., Monsefi, N., Herrero, A., Taylor, Cormac T., and von Kriegsheim, A. (2016). Substrate-Trapped Interactors of PHD3 and FIH Cluster in Distinct Signaling Pathways. Cell Reports 14, 2745–2760. 10.1016/j.celrep.2016.02.043.

81. Wooten, M.W. (1999). Function for NF-kB in neuronal survival: Regulation by atypical protein kinase C. J Neurosci Res 58, 607–611. 10.1002/(SICI)1097-4547(19991201)58:5<607::AID-JNR1>3.0.CO;2-M.

82. Tate, J.G., Bamford, S., Jubb, H.C., Sondka, Z., Beare, D.M., Bindal, N., Boutselakis, H., Cole, C.G., Creatore, C., Dawson, E., et al. (2018). COSMIC: the Catalogue Of Somatic Mutations In Cancer. Nucleic Acids Research 47, D941–D947. 10.1093/nar/gky1015.

83. Nesta, A.V., Tafur, D., and Beck, C.R. (2021). Hotspots of Human Mutation. Trends in Genetics 37, 717–729. 10.1016/j.tig.2020.10.003.

84. Mah, I.K., Soloff, R., Izuhara, A.K., Lakeland, D.L., Wang, C., and Mariani, F.V. (2016). Prkci is required for a non-autonomous signal that coordinates cell polarity during cavitation. Dev Biol 416, 82–97. 10.1016/j.ydbio.2016.06.002.

85. Chen, Z., Duan, Y., Wang, H., Tang, H., Wang, S., Wang, X., Liu, J., Fang, X., and Ouyang, K. (2021). Atypical protein kinase C is essential for embryonic vascular development in mice. genesis *59*, e23412. 10.1002/dvg.23412.

86. Seidl, S., Braun, U., Roos, N., Li, S., Lüdtke, T.H.W., Kispert, A., and Leitges, M. (2013). Phenotypical Analysis of Atypical PKCs In Vivo Function Display a Compensatory System at Mouse Embryonic Day 7.5. PLOS ONE 8, e62756. 10.1371/journal.pone.0062756.

